# Benchmarking of a multi-biomarker low-volume panel for Alzheimer’s Disease and related dementia research

**DOI:** 10.1101/2024.06.13.24308895

**Authors:** Laura Ibanez, Menghan Liu, Aleksandra Beric, Jigyasha Timsina, Pat Kholfeld, Kristy Bergmann, Joey Lowery, Nick Sykora, Brenda Sanchez-Montejo, Will Brock, John P. Budde, Randall J. Bateman, Nicolas Barthelemy, Suzanne E. Schindler, David M Holtzman, Tammie L. S. Benzinger, Chengjie Xiong, Rawan Tarawneh, Krista Moulder, John C. Morris, Yun Ju Sung, Carlos Cruchaga

**Affiliations:** Department of Psychiatry, Washington University School of Medicine; Department of Neurology, Washington University School of Medicine; NeuroGenomics and Informatics Center, Washington University School of Medicine; Hope Center for Neurologic Diseases, Washington University School of Medicine; The Tracy Family SILQ Center, Washington University School of Medicine; Knight Alzheimer Disease Research Center, Washington University School of Medicine; Department of Radiology, Washington University School of Medicine; Division of Biostatistics, Washington University School of Medicine; Department of Neurology, University of New Mexico School of Medicine; Department of Genetics, Washington University School of Medicine

**Author notes:** Co-corresponding authors Laura Ibanez, PhD, Carlos Cruchaga, PhD. Co-first authors (Laura Ibanez, Menghan Liu, Aleksandra Beric).

## Abstract

Alzheimer’s Disease (AD) biomarker measurement is key to aid in the diagnosis and prognosis of the disease. In the research setting, participant recruitment and retention and optimization of sample use, is one of the main challenges that observational studies face. Thus, obtaining accurate established biomarker measurements for stratification and maximizing use of the precious samples is key. Accurate technologies are currently available for established biomarkers, mainly immunoassays and immunoprecipitation liquid chromatography-mass spectrometry (IP-MS), and some of them are already being used in clinical settings. Although some immunoassays- and IP-MS based platforms provide multiplexing for several different coding proteins there is not a current platform that can measure all the stablished and emerging biomarkers in one run. The NUcleic acid Linked Immuno-Sandwich Assay (NULISA™) is a mid-throughput platform with antibody-based measurements with a sequencing output that requires 15µL of sample volume to measure more than 100 analytes, including those typically assayed for AD. Here we benchmarked and compared the AD-relevant biomarkers including in the NULISA against validated assays, in both CSF and plasma. Overall, we have found that CSF measures of Aß42/40, NfL, GFAP, and p-tau217 are highly correlated and have similar predictive performance when measured by immunoassay, mass-spectrometry or NULISA. In plasma, p-tau217 shows a performance similar to that reported with other technologies when predicting amyloidosis. Other established and exploratory biomarkers (total tau, p-tau181, NRGN, YKL40, sTREM2, VILIP1 among other) show a wide range of correlation values depending on the fluid and the platform. Our results indicate that the multiplexed immunoassay platform produces reliable results for established biomarkers in CSF that are useful in research settings, with the advantage of measuring additional novel biomarkers using minimal sample volume.

## Background

Alzheimer’s Disease (AD) is characterized by the pathological accumulation of misfolded amyloid beta (Aβ) plaques, and hyperphosphorylated tau tangles [1, 2]. Thus, most efforts have been focused on developing accurate measurements of amyloid and tau species to assist in the diagnosis and pathological monitoring of the disease. The amount and distribution of amyloid and tau pathology can be quantified in using Positron Emission Tomography (PET) imaging following injection of specific radiotracers, which is commonly used as reference standard for AD pathology in living individuals [3, 4]. However, the use of fluid biomarkers is more convenient, less invasive, and cost-effective. Levels of some CSF analytes are highly correlated with amyloid and tau pathology while the performance in plasma assays vary widely, with only some demonstrating high correlation with amyloid pathology [3, 5-8].

In both CSF and plasma, immunoprecipitation liquid chromatography-mass spectrometry (IP-MS) is the technology that has provided the highest correlation values with amyloid and tau PET and outperformed other assays in classification of pathologically positive *vs*. negative patients [9-11]. Especially, p-tau measurements that not only provide good correlation with cognitive decline but also outperform other measurements [9, 10, 12, 13], even when measured in plasma. Despite the good performance and the capability to measure many species of immunopurified protein, IP-MS multiplexing (measuring multiple single gene-coding proteins in one run) remains a challenge. Additionally, IP-MS requires relatively high volumes per analyte, specialized facilities, uses specialized machinery, and highly trained personnel which makes it difficult to use in a research setting. The need of bridging samples across measuring rounds is a limitation for both IP-MS and immunoassays, it increases the use of biofluid and the harmonization of values when performing longitudinal analyses its complex.

The use of immunoassays, despite having slightly poorer performance compared to IP-MS, is widespread. CSF assays (Aβ40, Aβ42, tau phosphorylated at position 181 (p-tau181) and more recently 217 (p-tau217)) show good correlation with Aβ brain deposits (as measured with amyloid PET) and acceptable discriminatory power to discern between amyloid positive and negative individuals [14, 15], especially when combined with *APOE* genotype and/or age [16-19]. IP-MS assays, especially for p-tau217 have also show very high predictive power to identify amyloid positive individuals based on imaging [8]. However, as the levels of some of the proteins are lower in plasma, immunoassays have presented with some performance challenges.

Besides Aβ40, Aβ42 and tau species, there are other biomarkers that are of interest including neurofilament light chain protein (NfL), neurogranin (NGRN), visinin like protein 1 (VILIP-1), soluble triggering receptor expressed on myeloid cells 2 (sTREM2), Chitinase 3 Like 1 (YKL40), and synaptosomal-associated protein 25 (SNAP25). NFL, and NGRN have been proposed as markers of neurodegeneration, a hallmark for AD and other neurodegenerative diseases [12]. Decreased levels of sTREM2, a marker of microglial activation and inflammatory changes, have been associated with increased rate of AD progression and is reported to be an endophenotype for AD [13–16]. Similarly, SNAP25, a member of the soluble N-ethylmaleimide-sensitive factor attachment protein receptors (SNARE) complex, has been proposed to be associated with decline in cognitive function resulting from synapse degradation, whereas variants and reduced levels of VILIP-1 have also been implicated in AD pathology [17–20]. Previous works have shown variable correlation values between measurements of these analytes assessed with high-throughput platforms such as SomaScan as compared with more classic immuno-assays [20, 21]. However, neither SomaScan nor Olink includes all the current relevant AD biomarkers. High and mid throughput technologies have been used to not only assess AD biomarkers, but also to perform discovery of new biomarkers and build predictive models [22-24].

Participant recruitment and retention along with sample acquisition, especially CSF via lumbar puncture, is one of the main challenges that observational studies face. Thus, maximizing use of the precious samples is key, especially in research settings. Volume is a minor limitation in a clinical setting. Input volume is highly dependent on the technology, but in general high and mid-throughput technologies require low volumes (55-100ul) to obtain thousands of measurements, which make them the most efficient for both volume and cost. There are several high throughput platforms available, including the relatively widely used Olink and SomaScan. Olink is an antibody-based technology that leverages proximity extension assays whereas SomaScan is aptamer based. However, in addition to the lack of robust measurements of established AD biomarkers, they present two additional disadvantages: (i) there is variable correlation between high throughput measures and the IP-MS or immunoassays values; [25] differences in correlation may be related to differences in the epitope measured, and differences in the binding efficiencies of the antibodies/aptamers; and (ii) most of the assays are not able to provide absolute quantifications values, which are typically used for medical interpretation. NUcleic acid Linked Immuno-Sandwich Assay (NULISA™) [26] is a mid-throughput platform that uses antibody-based measurements with a sequencing output. The main advantages are the low volume required (10 µL) combined with a high multiplexing capacity (more than 100 analytes) [26]. The central nervous system (CNS) panel includes traditional AD biomarkers, including Aβ40, Aβ42, p-tau181, and p-tau217 along with emerging and other AD biomarkers such as Aβ38, NFL, SNAP25, S100B, p-tau231, and p-tau217 among others.

Herein, we have benchmarked a new technology performing a head-to-head comparison of measures from the novel NULISA assay, IP-MS assays, and traditional immunoassays for Aβ40, Aβ42, Aβ42/40, total tau, p-tau181, and p-tau217 to assess utility in a research setting. For emerging biomarkers (NFL, NRGN, sTREM2, YKL40, SNAP25, GFAP, and VILIP1) we have compared NULISA to traditional immunoassays and high-throughput measurements. Finally, for other non-AD specific biomarkers we have compared NULISA to SomaScan and Olink.

## Material and Methods

### Participants

All research was performed in accordance with the protocols approved by the Institutional Review Board (IRB) at Washington University in Saint Louis.

We included 44 fasted CSF samples from 44 individuals and 44 unfasted plasma samples from 44 individuals from the Memory and Aging Project (MAP) at the Charles F. and Joanne Knight Alzheimer Disease Research Center (Knight-ADRC) at Washington University in Saint Louis with biomarker data measured available in CSF and/or plasma using reference and high-throughput assessments. Due to the benchmarking characteristics of this study, the inclusion criterion was data availability, no clinical criteria were used. Despite the presence of some individuals being present in both CSF and plasma samples (N=69), CSF and plasma samples/visits were not matched, and therefore represent different cohorts (Table 1) for the purpose of this study. As part of the regular study protocol, specimen collection, and imaging assessments are obtained at least every two years. CSF samples are collected close to the clinical visit at the Knight-ADRC MAP clinic after an overnight fast along with fasted and non-fasted plasma. CSF and plasma are immediately processed and stored at -80°C until used. Amyloid scans (obtained within six months before or after the biofluid collection) obtained and processed as previously described [27] were available for 43 of the 44 CSF samples and 39 out of 44 plasma samples.

**Table 1.** Demographic characteristics of the Knight-ADRC participants included in each of the two cohorts assessed in the present study.

|  | **CSF Cohort** | **Plasma Cohort** |
| --- | --- | --- |
| **Sample Size (N)** | 44 | 44 |
| **Sex (N, % Females)** | 31 (70.45) | 19 (43.18) |
| **Age at Draw\*** | 65 (50, 91) | 71 (50, 91) |
| **Non-Hispanic White (N, %)** | 42 (95.45) | 42 (95.45) |
| **APOε4 carriers<sup>#</sup> (N, %)** | 16 (36.36) | 24 (54.55) |
| **PET Availability (N, %)** | 43 (97.73) | 39 (88.64) |
| **Positive Amyloidosis<sup>&</sup> (N, %)** | 14 (31.82) | 20 (45.45) |
| **Clinical AD Cases (N, %)** | 6 (13.64) | 9 (20.45) |

\*mean (maximum, minimum); <sup>#</sup>Carriers of at least one APOε4 allele;
<sup>&</sup>Positivity calculated using the normalized amyloid PET centiloid and the GMM method; CSF=Cerebrospinal Fluid; PET= Positron emission tomography; AD=Alzheimer's Disease

For those participants with PET scans available (N=43 for CSF and N=39 for plasma samples), cortical Aß depositions were quantified using ^11^C Pittsburgh Compound-B (PiB) PET and ^18^F-AV-1451 (florbetapir) PET. Individuals were classified as amyloid positive (N=14 for CSF samples, and N=20 for plasma samples) or negative (N=29 for CSF samples, and N=19 for plasma samples) based on imaging quantification. Given the use of two compounds, centiloids were standardized using Z-scores and positivity was assessed using a gaussian mixture model (GMM) data driven approach as previously described[21]. Additionally, all participants were stratified as AD cases (N=6 for CSF, and N=9 for plasma) or controls (N=35 for CSF and N=31 for plasma) based on clinical dementia rating (CDR^®^) and clinical diagnosis of AD. We were unable to classify three CSF samples and four plasma samples using this criterion.

### NUcleic acid Linked Immuno-Sandwich Assay (NULISA™) Measurements

NULISA [26] is a novel technology based on a mechanism of dual capture and release. This approach minimizes assay background and increases sensitivity at the attomole level. Briefly, DNA conjugated antibodies form immunocomplexes with the target protein in solution, the immunocomplex is purified sequentially, first with an oligo-dT beads binding to the polyA sequence attached to the capture antibody, and then with streptavidin beads binding to the biotin on the detection antibody. Then, the DNA reporter is generated by proximity ligation and amplified by PCR to generate next-generation sequencing libraries. The DNA oligonucleotide contains the barcode to de-multiplex targeted protein and sample identification. This technology has the capability of measuring more than 200 proteins in the same 10µL of sample. To measure the samples included in this study we have utilized the NULISAseq CNS Disease Panel 120. Concentration is reported in NULISA Protein Quantification (NPQ) units. NPQ units are the result of normalizing the sequence quantification counts by intraplate and intensity variability and log_2_-transformed to approximate normality.

### Immunoassay and Mass Spectrometry Measurements

All measurement were obtained from the Knight-ADRC Biomarker Core. Briefly, CSF Aβ40, Aβ42, total tau, and p-tau181 measurements were performed using a Lumipulse G1200 Chemiluminescent Enzyme Immunoassay Assay platform as previously described [3, 20, 21]. In contrast, plasma Aβ40 and Aβ42 levels were measured using mass spectrometry [5, 8] along with CSF p-tau217 [12, 28]. Only IP-MS concentration values have been retained for analyses. Other emerging AD biomarkers were quantified using a variety of immunoassays. CSF NfL was measured using commercially available immunoassays kits (Uman Diagnostics AB, Umea, Sweden) as previously described [29-31], whereas plasma levels were assessed using Quanterix[32]. CSF NRGN, SNAP25, and VILIP1 were performed using Single Molecule Counting (SMC) technology (EMD Millipore, Burlington, MA) as previously described [33-36]. YKL40 CSF concentration were measured using the MicroVue YKL40 EIA plate-based enzyme immunoassay (Quidel, San Diego, CA) for serum and plasma kit. We followed a previously described modified version of the protocol that allows quantification in CSF fluid. Briefly, samples were diluted threefold prior to assay, and washes included five cycles instead of four. Remaining steps were followed as indicated by the manufacturer [37]. Finally, sTREM2 was measured using an in-house immunoassay [38-40] and plasma GFAP was measured using the ultra-sensitive single-molecule array (Simoa, Quanterix) [41].

### High-Throughput Measurements

High-throughput aptamer-based protein measurements using SomaScan 7K were available for all plasma and CSF samples included in this comparison [42, 43]. In short, modified single-stranded DNA aptamers will specifically bind the targeted protein. After washing and processing, the aptamers are quantified by a DNA microarray. Quantification of protein concentrations is done using relative fluorescent units (RFU). Prior to analysis relative quantifications undergo stringent quality control, described elsewhere [23, 24]. Even though the full panel includes around seven thousand aptamers, we only included in here the ones that overlap with the NULISAseq CNS Panel and pass quality control in both platforms.

Similarly, 43 CSF samples were also measured using Olink Explore HT1. No Plasma samples were measured using Olink. This technology allows to measure thousands of proteins in a multiplex manner using Proximity Extension Assays (PEA) and Illumina Next Generation Sequencing using minimal sample volumes. Relative protein quantification is done via Normalized Protein eXpression (NPX) values that are already adjusted for intra and inter assay variability and log_2_ transformed to approximate normal distribution.

### Statistical Analyses

All analyses were performed in R Project for Statistical Computing, including all the available data for each analyte, individual, and fluid. All the raw values corresponding to protein concentration and relative fluorescent units (RFU) from SomaScan were log_10_ transformed to approximate the normal distribution. NPQ values from NULISA are already log_2_ transformed and no transformation was used. Then, head-to-head comparisons were performed using Spearman correlations.

For differential abundance analyses we used case control classification along with quantitative imaging amyloid burden. The centiloid values were log_10_ transformed to approximate normality and used as phenotype in a regression. For both imaging and case/control status in plasma and CSF, the *lm* function from the *stats* R package was used to determine association. All analysis were adjusted by age and sex. P-values were FDR corrected to account for multiple testing.

Predictive performance comparison across measurement technologies were performed by using receiver operating characteristic (ROC) curves and calculating the area under the ROC curves (AUC). Then, we used multivariate logistic regression using normalized protein values to predict amyloid positivity. Additionally, we also included age and sex as covariates to account for baseline risk to evaluate if there was improvement in the prediction. Due to the limited sample size, no statistical test was used to formally compare performance.

## Results

### Established and emerging CSF AD Biomarkers

We compared the biomarker concentration of all the established AD biomarkers (Aβ40, Aβ42, Aβ42/40 ratio, total tau, and p-tau181) measured using NULISA (NPQ values) and Lumipulse (pg/mL; Figure 1A, Table S1). We observed that Aβ40 measured with Lumipulse and NULISA was highly correlated (ρ=0.73 [0.53-0.84], p=2.38× 10^-08^; Figure S1A) along with Aβ42 (ρ=0.84 [0.70-0.92], p=7.18× 10^-13^; Figure S1B). The Aβ42/40 ratio correlation was higher that the Aβ species individually, with ρ=0.91 ([0.81-0.95], p=1.04×10^-17^; Figure S1C). We also tested the correlation between Aβ42/40 ratio and amyloid PET centiloid. The ratio obtained from Lumipulse showed a correlation of ρ=0.78 [0.59-0.89], (p=7.51×10^-10^, Figure S2A), similar to the one obtained from NULISA ρ=0.74 [0.53-0.85] (p=1.81×10^-08^, Figure S2B). Total tau Lumipulse concentrations were correlated with NULISA (ρ=0.83 [0.63-0.95], p=2.96×10^-12^; Figure S1D). Similarly, p-tau181 Lumipulse values also correlated with NULISA (NPQ values ρ=0.98 [0.95-0.99], p=3.07×10^-33^; Figure S1E). IP-MS was used to measure CSF p-tau217 concentration (pg/mL), that also showed high correlation with NULISA quantification (ρ=0.96 [0.92-0.98], p=1.60×10^-25^; Figure S1F). Additionally, CSF p-tau217 concentration was correlated with centiloid values when measured with IP-MS (ρ=0.70 [0.49-0.83], p=2.00×10^-07^; Figure S2C), and NULISA (ρ=0.75 [0.56-0.85], p=8.55×10^-09^; Figure S2D).

**Figure 1.**
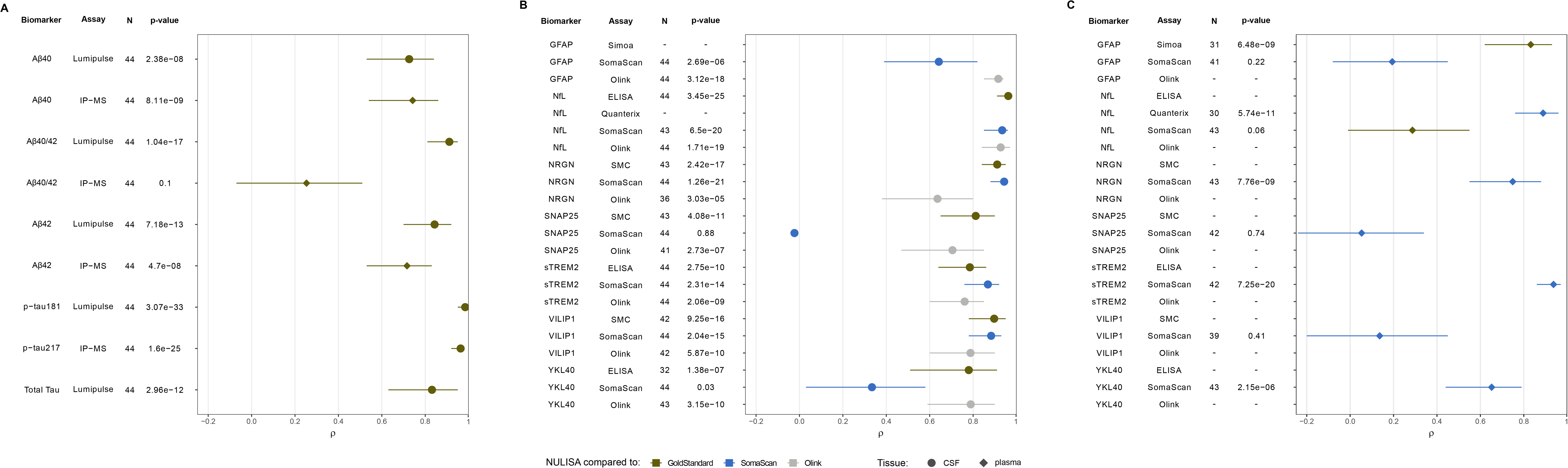
Correlations between NULISA and other quantification methods for **A**. established AD biomarkers in CSF and plasma using gold-standard assessments, **B**. emerging biomarkers in CSF, and **C**. emerging biomarkers in Plasma. In green are indicated the comparison between NULISA and gold-standard measurements, in blude the comparisons to SomaScan, and in grey the comparisons to Olink. Circles indicate correlations in CSF whereas diamonds indicate correlations in plasma.

Then, we analyzed markers of neurodegeneration, NfL and NRGN (Figure 1B, Table S1). NfL NULISA strongly correlated with immunoassay (ρ=0.96 [0.91-0.98]; p=3.45×10^-25^; Figure S3A), SomaScan-based measures (ρ=0.93 [0.85-0.96]; p=6.50×10^-20^; Figure S3), and Olink (ρ=0.93 [0.84-0.97]; p= 1.71×10^-19^; Figure S3C). High correlations were also found for NULISA NRGN when comparing with SomaScan (ρ=0.94 [0.88-0.96]; p=1.26×10^-21^; Figure S3D), as well as SMC (ρ=0.91 [0.84-0.95]; p=2.42×10^-17^; Figure S3E), but lower, yet significant with Olink (ρ=0.64 [0.38-0.80]; p=3.03×10^-05^; Figures S3F).

CSF sTREM2 levels, a marker of microglia activation and inflammation, were available in the four platforms (Figure 1B, Table S1), with an ρ=0.79 ([0.64-0.86], p=2.75×10^-10^) when comparing immuno-assay and NULISA (Figure S4A) and ρ=0.87 ([0.76-0.92], p=2.31×10^-14^) when comparing SomaScan and NULISA quantifications (Figure S4B). Olink measurements were also significantly correlated with NULISA quantification (ρ=0.76 [0.60-0.85], p=2.06×10^-09^; Figure S4C). YKL40, another biomarker of neuroinflammation, measurements using the immunoassay also showed significant correlation with NULISA values (ρ=0.78 [0.51-0.91]; p=1.38×10^-07^; Figures 1B, S4D, Table S1), but not with those from SomaScan (ρ=0.33 [0.03-0.58], p=0.03; Figure 1S4E). However, Olink measurements were highly correlated with those from NULISA (ρ=0.79 [0.59-0.90], p=3.15×10^-10^; Figure S4F).

The biomarker for neuronal injury VILIP1 NULISA values showed high correlation across all three platforms (Figure 1B, Table S1). SMC quantification was highly correlated with NULISA relative quantification (ρ=0.90 [0.78-0.95], p=9.26×10^-16^; Figure S5A), with SomaScan (ρ=0.88 [0.78-0.93], p=2.04×10^-15^; Figure S5B), and Olink (ρ=0.79 [0.60-0.90], p=5.87×10^-10^; Figure S5C). SNAP25, a biomarker of synapse loss, also showed correlation between SMC and NULISA (ρ=0.81 [0.65-0.90], p=4.08×10^-11^; Figures 1B, S5D, Table S1), Olink and NULISA (ρ=0.71 [0.47-0.85], p=2.73×10^-07^; Figure S5E) but not between NULISA and SomaScan (ρ=-0.02 [-0.35-0.27], p=0.88; FigureS5F) as reported before [20]. CSF GFAP measurement, a marker of astrogliosis was available in NULISA and SomaScan (Figure 1B, Table S1), with a moderate but significant correlation (ρ=0.64 [0.39-0.82], p=2.69×10^-06^; Figure S5G), and Olink (ρ=0.92 [0.85-0.94], p=3.12×10^-18^; Figure S5H).

### Plasma AD traditional and emerging biomarkers

To assess the performance of NULISA assessments in plasma, we performed similar analyses as those described for CSF (Figure 1A, Table S1). IP-MS Aβ40 concentration was significantly correlated with NULISA relative quantification (ρ=0.74 [0.54-0.86], p=8.11×10^-09^; Figure S6A), along with that of Aβ42 (ρ=0.72 [0.53-0.83], p=4.70×10^-08^; Figure S6B). However, the Aβ42/40 ratio showed poor correlation between IP-MS and NPQ quantifications (ρ=0.25 [-0.07-0.51], p=0.10; Figure S6C). Along the same lines, the correlation of Aβ42/40 obtained from IP-MS measurements showed higher correlation (ρ=0.64 [0.40-0.82], p=1.20×10^-05^; Figure S2F) than the one from NULISA assessments (ρ=0.37 [0.04-0.63], p=0.02; Figure S2G). Plasma p-tau181 and p-tau217 were not available for head-to-head comparison. We did however observe a highly significant correlation between p-tau217 and amyloid PET centiloid (ρ=0.72 [0.55-0.85], p=1.87×10^-07^; Figure S2H). NfL plasma measurements measured using Quanterix were highly correlated when compared to those assessed with NULISA (ρ=0.89 [0.76-0.94], p=5.74×10^-11^; Figures 1C, S6D, Table S1) but not with SomaScan (ρ=0.29 [-0.01-0.55], p=0.06; Figure S6E). Plasma sTREM2 measurements with SomaScan were highly correlated with NULISA (ρ=0.94 [0.86-0.97], p=7.24×10^-20^; Figures 1C, S6F, Table S1), whereas NRGN SomaScan values showed a moderate but significant correlation with NULISA (ρ=0.75 [0.55-0.88], p=7.76×10^-09^; Figures 1C, S6G, Table S1). The YKL40 correlation between SomaScan plasma quantification and NULISA was significant (ρ=0.65 [0.44-0.79], p=2.15×10^-06^; Figures 1C, S6H, Table S1). No correlation was observed between SomaScan and NULISA measurements for SNAP25 (ρ=0.05 [-0.24-0.34], p=0.74; Figure 1C, Table S1) or VILIP1 (ρ=0.14 [-0.20-0.45], p=0.41; Figure 1C, Table S1). Finally, plasma GFAP showed high correlation between Simoa and NULISA (ρ=0.83 [0.62-0.93], p=6.48×10^-09^; Figures 1C, S6I, Table S1), but not between SomaScan and NULISA (ρ=0.19 [-0.08-0.45], p=0.22; Figure 1C, Table S1).

### Comparison of NULISA with High Throughput measurements for other proteins

Similar to traditional biomarkers, we assessed if the measurements of the aptamer-multiplex platform SomaScan and the PEA-based platform Olink, were comparable to those quantified using the antibody-multiplexed platform quantifications NULISA (Table S2). We observed a wide range of correlation values for CSF (Figures S7A and S7B). SomaScan had 102 analytes in common with NULISA. The best correlations were observed for proteins such as BASP1 (Brain Abundant Membrane Attached Signal Protein 1, related to apoptotic mechanisms), NPTX2 (Neuronal Pentraxin II), or CXCL8 (Interleukin 8) with ρ values close to one, whereas others like IL10 (Interleukin 10), CD40L (CD40 ligand), or TNF (Tumor Necrosis Factor) showed the worst correlations (Table S2). There were 97 CSF proteins measured using Olink and NULISA. Overall, we observed a lower correlation with NULISA than SomaScan (Table S2). Top correlations between NULISA and Olink were observed for emerging biomarkers NFL, YKL40, and GFAP, with ρ>0.90 and discussed in the previous section. None of the other quantifications show ρ>0.90, MDH1 (Malate Dehydrogenase 1) showed the highest correlation (ρ=0.87 [0.73-0.93], p=2.62×10^-^ _13_), followed by NPTX2 (ρ=0.86 [0.74-0.93], p=7.08×10^-14^). Non correlated measurements included BDNF (Brain-Derived Neurotrophic Factor), NPTXR (Neuronal Pentraxin Receptor) or CD40L among others. Differences in the targeted epitopes or in the sensitivity between assays might explain the wide range of correlation values observed, as previously described [26, 44].

In plasma, 95 protein measurements were available in NULISA and SomaScan. No measurement with Olink were available. Similar to CSF, the correlation ranges were very wide (Table S2, Figure S7C). Top correlated measurements include alpha synuclein, sTREM2, BDNF, and CRP (C-Reactive Protein), while the less correlated included established AD biomarkers such as p-tau217, p-tau231, and p-tau181.

### NULISA biomarker levels association against amyloid imaging and clinical status

Next, we performed a differential abundance analysis using logistic regression to confirm the association with amyloid PET and clinical status for the most relevant proteins included in the NULISA CNS panel.

PET amyloid imaging was available for the 43 individuals with CSF measurements. We found the levels of ten CSF proteins significantly associated with the harmonized centiloid amyloid imaging measurements [21] (Figure S8A, Table S3). Of those, p-tau-217 (beta= 0.91 [0.68,1.14], p=2.45×10^-09^), p-tau231 (beta= 0.73 [0.52,0.94], p=5.30×10^-08^), p-tau-181 (beta=0.46 [0.28,0.64], p=1.44×10^-05^), and total tau (Microtubule Associated Protein Tau (MAPT)-beta=0.35 [0.17,0.54], p=6.34×10^-04^) showed the strongest association and remained significant after multiple test correction and adjusting by sex and age. Several other proteins were nominally associated with amyloid PET quantification (Aβ42, VILIP1, IL6R, YWHAG and TIMP3.

We also performed similar analyses in plasma (N=39) and found 18 proteins significantly associated with amyloid accumulation measured with PET (Figure S8B, Table S3). The levels of p-tau217 (beta=0.31 [0.23,0.39], p=6.22×10^-09^) showed the strongest association with amyloid accumulation and remained significant after FDR correction followed by p-tau231 (beta=0.25 [0.10,0.40], p=2.33×10^-03^), GFAP (beta=0.25 [0.09,0.41], p=0.004) and Aβ42 (beta=-0.17 [-0.31,-0.03], p=0.02). The remaining associations were related to the inflammatory pathway: CCL13; CCL26, CXCL8 IGFBP7, TIMP3, IL7, IL1B, SQSTM, BDNF, MDH1, FGF2, MME and HBA1.

When using clinical case-control classification for the differential abundance analyses, the results changed drastically, with a clear reduction in statistical power with six and nine proteins nominally significant in CSF and plasma respectively (Table S3). Of note, the comparison included six clinical AD cases for CSF, and nine in plasma. In CSF, IL1β was the protein with the strongest association (beta=-0.12 [-0.19,-0.05], p=1.76×10^-03^). We were also able to detect Aβ42 (beta=-1.20 [-2.20,-0.20], p=0.02; Figure S8C) even with the unbalanced population studied. In plasma the strongest associations observed for NfL (beta=0.90 [0.45,1.35], p=3.99×10^-04^) and p-tau217 (beta=0.49 [0.12,0.87], p=0.01; Figure S8D).

### Amyloid positivity predictive performance across platforms and tissues

We assessed the performance of each platform and protein to predict amyloid positivity assessed by PET for both CSF and plasma measurements as standalone measurement and adding age and sex at birth as covariates (Table S4). In CSF, the Aβ42/40 ratio showed an AUC of 0.91 [0.82-1.00] for immunoassay and 0.94 [0.88-1.00] for NULISA-based measurements when including sex and age (Figure 2A). Analogously, no performance differences were found for p-tau217 between IP-MS (AUC=0.92 [0.84-1.00]) and NULISA (AUC=0.93 [0.86-1.00]; Figure 2A). similarly, p-tau181 showed an AUC of 0.89 [0.79-0.99] and 0.90 [0.73-0.96] when measured using immunoassay techniques and NULISA respectively (Figure 2A). The AUC values for Total tau were very close when measured with NULISA (AUC=0.85 [0.73-0.96]) or with immunoassay (AUC=0.83 [0.71-0.95]; Figure 2A). NfL showed limited discriminatory performance regardless of the technology (Table 3). Finally, NULISA p-tau231 showed similar classification performance (AUC=0.92 [0.84-1.00]; Figure 2A) than that previously reported in the literature [45].

**Figure 2.**
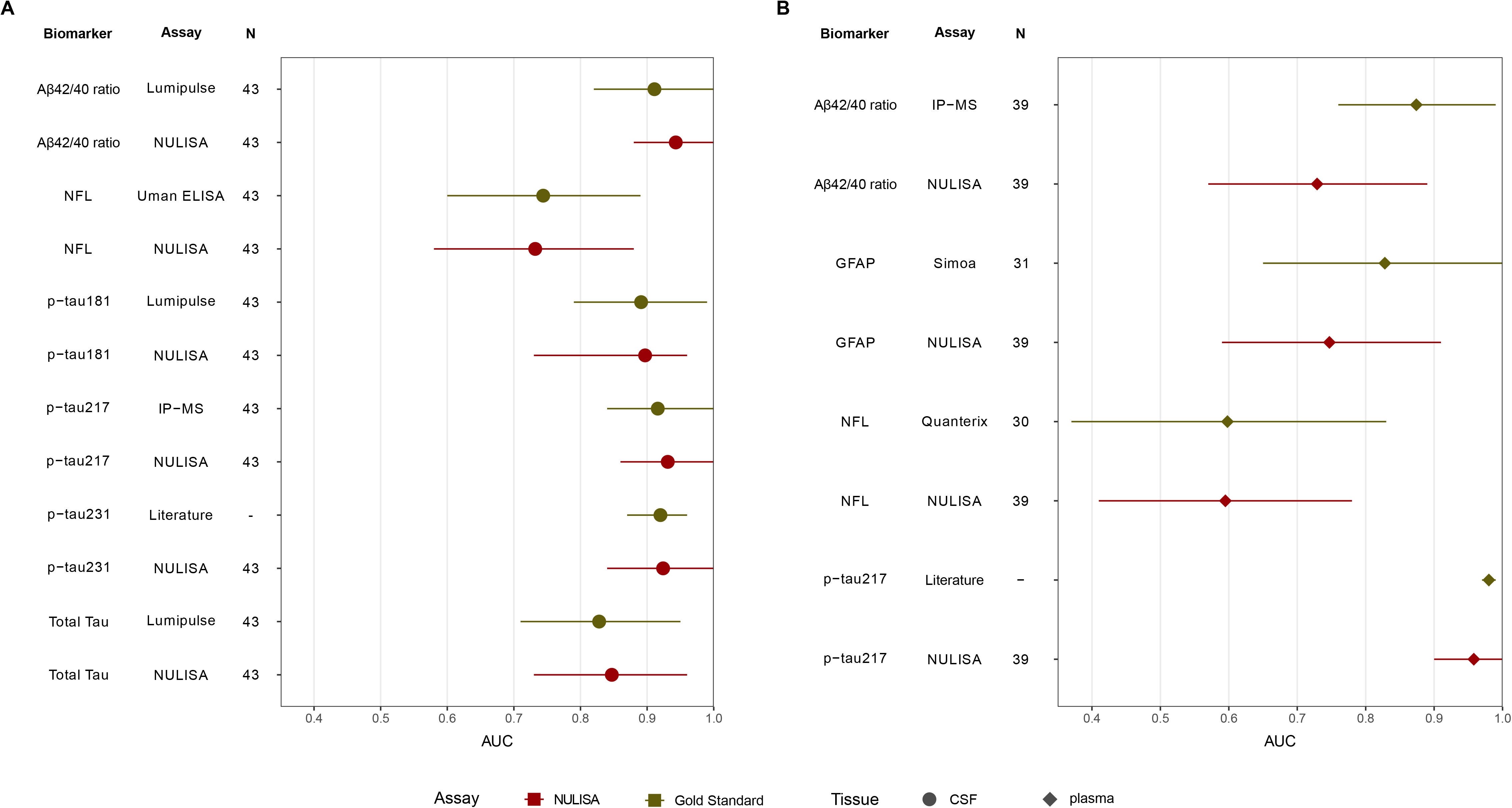
Performance of established and emerging biomarkers to classify brain amyloid positivity (based on Pittsburgh Compound-B (PiB) Positron emission tomography (PET) quantifications) expressed as Area Under the Receiver Operating Characteristic Curve (AUC) in **A**. CSF (circle), and **B**. Plasma (diamond). Red indicates AUC for NULISA, whereas green indicates the AUC for the gold standard method.

In plasma, Aβ42/40 ratio show lower performance in NULISA (AUC=0.73 [0.57-0.89]; Figure 2B) compared to IP-MS (AUC=0.87 [0.76-0.99]; Figure 2B). Despite no comparison being available, p-tau217 measured by NULISA had similar predictive power (AUC=0.96 [0.90-1.00]; Figure 2B) than that reported previously [46]. The AUC for NfL was similar for both immunoassay and NULISA, regardless of the addition of covariates (AUC=0.60 [0.37-0.83] and AUC=0.60 [0.41-0.78] for immunoassay and NULISA respectively; Figure 2B), whereas for GFAP the predictive power of the immunoassay (AUC=0.83 [0.65-1.00]; Figure 2B) was higher than with NULISA (AUC=0.75 [0.59-0.91]; Figure 2B), in both cases when combined with age and sex.

## Discussion

As the research community describes novel and better biomarkers, technology also advances to enable multiplex accurate measurements that use low sample volume. Using multiplex assays streamlines the integration of measurements from the same visit and reduce sample swap errors due to the processing of one sample per patient to obtain several quantifications, instead of processing several samples to obtain individual measurements than need to be matched. Low volume assays facilitate bridging and extends the use of each biological fluid. Overall, multiplex measurements are less prone to error, and more cost-efficient from the hands on-time and sample usage perspective. NULISA is a novel technology lacking a head-to-head comparison in the context of neurodegeneration-related biomarkers. Here, we have compared the NULISA quantifications (NPQ values) with those obtained using the gold-standard method (immunoassay or mass spectrometry-based) and high-throughput (SomaScan and Olink) quantifications to assess the accuracy and predictive power of this novel measurements in both CSF and plasma.

Overall, we have found that in CSF the NULISA values for the established biomarkers using well-validated gold standard assays (Lumipulse Aß42, Aß40, Aß42/40 ratio, total tau, p-tau181, and IP-MS p-tau217) are highly correlated and show similar performance in classifying amyloid PET status. In plasma, p-tau217 shows a similar AUC that the one reported in the literature [10, 46], as well as ptau231 [45], but the predictive power for Aß40/42 is significantly lower than established assays. These differences between CSF and plasma could be explained by the protocol optimization of IP-MS and immunoassay based on fluid. NULISA uses the same panel of antibodies and protocol regardless of tissue. Thus, it is plausible to think that the epitopes captured in CSF and plasma are optimized for immunoassay, but not for NULISA. Optimization of the NULISA CNS panel for plasma might improve the results reported in here.

When focusing on the emerging biomarkers, NULISA quantifications showed significant correlations for all the AD-relevant biomarkers such as sTREM2, GFAP, YKL40 when those were quantified using reference standard technologies. With high-throughput technologies the results are more variable, especially for those assays known to be non-optimal in the high-throughput platform such as YKL40 or SNAP25 in SomaScan. These findings suggest that NULISA can aid in the assessment of all AD-related biomarkers in multiplex manner for the first time.

Due to the inclusion of more than 100 analytes in the CNS panel, and the aim to assess the panel in a research setting, we performed differential abundance analysis to test if we were able to identify previously associated AD proteins, and potentially novel ones. The top protein in both CSF and plasma was p-tau217, already described to be an AD biomarker [28], closely followed by p-tau231. While in CSF we identified p-tau181, MAPT, Aß42 and NGRN, already associated and known biomarkers for AD, only p-tau231 and GFAP were identified in plasma. All findings were in the correct direction of effect, supporting that correctly powered samples assessed using NULISA technology have the power to evaluate established AD biomarkers, measure AD-related processes such as neuronal injury or inflammation, and identify novel differentially accumulated proteins in a research setting.

This study has several limitations. The main one is the limited sample size used for the benchmarking, which could lead to wide confidence intervals for the correlations and the AUC calculations. For the same reason, we have not applied formal statistical comparison when calculating AUCs. However, the aim of this study was to assess the utility of the NULISA platform in research settings and assess the accuracy of its measures compared to gold standard and established assays measurements. Each assay performs within a range of concentrations, thus formal assay comparison in larger sample sizes with a wide range of concentrations could help assess if the lack of correlation, of special interest in plasma Aβ species quantification is due to lack of optimization or limit of detection. Overall, and despite the limitations, this work supports the use of NULISA quantification for research purposes, and evidence that the NPQ values have biological meaning and can be used to ask scientific questions. They provide accurate measurements and optimizes the use of precious biofluids. Similar analyses in larger samples sizes, multi-site studies, and diverse cohorts are needed to further validate its use in research or clinical settings.

In conclusion, this study reports for the first time the correlation values between established AD biomarker measurements assessed using the gold standard and a novel multiplexed assay. Given the high correlation in CSF and plasma, the minimal sample requirement, and the multiplexing capacity of NULISA, these assays have the potential of revolutionizing the scientific field by providing highly efficient measurements, saving precious sample and thus providing the means for more discoveries.

## Data Availability

All data produced in the present study are available upon reasonable request to the authors

## Acknowledgements

We thank all the participants and their families, as well as the many involved institutions and their staff.

This work was supported by grants from the National Institutes of Health (R01AG044546 (CC), P01AG003991(CC, JCM), RF1AG053303 (CC), RF1AG058501 (CC), U01AG058922 (CC), K99/R00-AG062723 (LI), RF1AG074007 (YJS), R01AG070941 (SES)), the Chan Zuckerberg Initiative (CZI), the Michael J. Fox Foundation (LI, CC), the Department of Defense (LI-W81XWH2010849), the Alzheimer’s Association Zenith Fellows Award (ZEN-22-848604, awarded to CC), the Alzheimer’s Drug Discovery Foundation (LI), Bright Focus Foundation (LI), and an Anonymous foundation.

The recruitment and clinical characterization of research participants at Washington University were supported by NIH P30AG066444 (JCM), P01AG03991(JCM), and P01AG026276(JCM).

This work was supported by access to equipment made possible by the Hope Center for Neurological Disorders, the Neurogenomics and Informatics Center (NGI: https://neurogenomics.wustl.edu/)and the Departments of Neurology and Psychiatry at Washington University School of Medicine.

## References

1. Hardy J, Allsop D: Amyloid deposition as the central event in the aetiology of Alzheimer’s disease. Trends in pharmacological sciences 1991, 12(10):383–388.

2. Selkoe DJ: The molecular pathology of Alzheimer’s disease. Neuron 1991, 6(4):487–498.

3. Wisch JK, Gordon BA, Boerwinkle AH, Luckett PH, Bollinger JG, Ovod V, Li Y, Henson RL, West T, Meyer MR et al: Predicting continuous amyloid PET values with CSF and plasma Abeta42/Abeta40. Alzheimer’s & dementia 2023, 15(1):e12405.

4. Hansson O, Edelmayer RM, Boxer AL, Carrillo MC, Mielke MM, Rabinovici GD, Salloway S, Sperling R, Zetterberg H, Teunissen CE: The Alzheimer’s Association appropriate use recommendations for blood biomarkers in Alzheimer’s disease. Alzheimer’s & dementia : the journal of the Alzheimer’s Association 2022, 18(12):2669–2686.

5. Schindler SE, Bollinger JG, Ovod V, Mawuenyega KG, Li Y, Gordon BA, Holtzman DM, Morris JC, Benzinger TLS, Xiong C et al: High-precision plasma beta-amyloid 42/40 predicts current and future brain amyloidosis. Neurology 2019, 93(17):e1647–e1659.

6. Brand AL, Lawler PE, Bollinger JG, Li Y, Schindler SE, Li M, Lopez S, Ovod V, Nakamura A, Shaw LM et al: The performance of plasma amyloid beta measurements in identifying amyloid plaques in Alzheimer’s disease: a literature review. Alzheimer’s research & therapy 2022, 14(1):195.

7. Li Y, Schindler SE, Bollinger JG, Ovod V, Mawuenyega KG, Weiner MW, Shaw LM, Masters CL, Fowler CJ, Trojanowski JQ et al: Validation of Plasma Amyloid-beta 42/40 for Detecting Alzheimer Disease Amyloid Plaques. Neurology 2022, 98(7):e688–e699.

8. Barthelemy NR, Salvado G, Schindler SE, He Y, Janelidze S, Collij LE, Saef B, Henson RL, Chen CD, Gordon BA et al: Highly accurate blood test for Alzheimer’s disease is similar or superior to clinical cerebrospinal fluid tests. Nat Med 2024, 30(4):1085–1095.

9. Salvado G, Horie K, Barthelemy NR, Vogel JW, Binette AP, Chen CD, Aschenbrenner AJ, Gordon BA, Benzinger TLS, Holtzman DM et al: Novel CSF tau biomarkers can be used for disease staging of sporadic Alzheimer’s disease. medRxiv 2023.

10. Janelidze S, Bali D, Ashton NJ, Barthelemy NR, Vanbrabant J, Stoops E, Vanmechelen E, He Y, Dolado AO, Triana-Baltzer G et al: Head-to-head comparison of 10 plasma phospho-tau assays in prodromal Alzheimer’s disease. Brain : a journal of neurology 2023, 146(4):1592–1601.

11. Janelidze S, Teunissen CE, Zetterberg H, Allue JA, Sarasa L, Eichenlaub U, Bittner T, Ovod V, Verberk IMW, Toba K et al: Head-to-Head Comparison of 8 Plasma Amyloid-beta 42/40 Assays in Alzheimer Disease. JAMA neurology 2021, 78(11):1375–1382.

12. Barthelemy NR, Li Y, Joseph-Mathurin N, Gordon BA, Hassenstab J, Benzinger TLS, Buckles V, Fagan AM, Perrin RJ, Goate AM et al: A soluble phosphorylated tau signature links tau, amyloid and the evolution of stages of dominantly inherited Alzheimer’s disease. Nat Med 2020, 26(3):398–407.

13. Horie K, Salvado G, Barthelemy NR, Janelidze S, Li Y, He Y, Saef B, Chen CD, Jiang H, Strandberg O et al: CSF MTBR-tau243 is a specific biomarker of tau tangle pathology in Alzheimer’s disease. Nat Med 2023, 29(8):1954–1963.

14. Fagan AM, Mintun MA, Mach RH, Lee SY, Dence CS, Shah AR, LaRossa GN, Spinner ML, Klunk WE, Mathis CA et al: Inverse relation between in vivo amyloid imaging load and cerebrospinal fluid Abeta42 in humans. Ann Neurol 2006, 59(3):512–519.

15. Fagan AM, Roe CM, Xiong C, Mintun MA, Morris JC, Holtzman DM: Cerebrospinal fluid tau/beta-amyloid(42) ratio as a prediction of cognitive decline in nondemented older adults. Arch Neurol 2007, 64(3):343–349.

16. Fagan AM, Watson M, Parsadanian M, Bales KR, Paul SM, Holtzman DM: Human and murine ApoE markedly alters A beta metabolism before and after plaque formation in a mouse model of Alzheimer’s disease. Neurobiol Dis 2002, 9(3):305–318.

17. Cruchaga C, Kauwe JS, Harari O, Jin SC, Cai Y, Karch CM, Benitez BA, Jeng AT, Skorupa T, Carrell D et al: GWAS of cerebrospinal fluid tau levels identifies risk variants for Alzheimer’s disease. Neuron 2013, 78(2):256–268.

18. Cruchaga C, Kauwe JS, Mayo K, Spiegel N, Bertelsen S, Nowotny P, Shah AR, Abraham R, Hollingworth P, Harold D et al: SNPs associated with cerebrospinal fluid phospho-tau levels influence rate of decline in Alzheimer’s disease. PLoS genetics 2010, 6(9):e1001101.

19. Cruchaga C, Kauwe JS, Nowotny P, Bales K, Pickering EH, Mayo K, Bertelsen S, Hinrichs A, Alzheimer’s Disease Neuroimaging I, Fagan AM et al: Cerebrospinal fluid APOE levels: an endophenotype for genetic studies for Alzheimer’s disease. Hum Mol Genet 2012, 21(20):4558–4571.

20. Timsina J, Gomez-Fonseca D, Wang L, Do A, Western D, Alvarez I, Aguilar M, Pastor P, Henson RL, Herries E et al: Comparative Analysis of Alzheimer’s Disease Cerebrospinal Fluid Biomarkers Measurement by Multiplex SOMAscan Platform and Immunoassay-Based Approach. J Alzheimers Dis 2022.

21. Timsina J, Ali M, Do A, Wang L, Western D, Sung YJ, Cruchaga C: Harmonization of CSF and imaging biomarkers in Alzheimer’s disease: Need and practical applications for genetics studies and preclinical classification. Neurobiol Dis 2023, 190:106373.

22. Johnson ECB, Bian S, Haque RU, Carter EK, Watson CM, Gordon BA, Ping L, Duong DM, Epstein MP, McDade E et al: Cerebrospinal fluid proteomics define the natural history of autosomal dominant Alzheimer’s disease. Nat Med 2023, 29(8):1979–1988.

23. Yang C, Farias FHG, Ibanez L, Suhy A, Sadler B, Fernandez MV, Wang F, Bradley JL, Eiffert B, Bahena JA et al: Genomic atlas of the proteome from brain, CSF and plasma prioritizes proteins implicated in neurological disorders. Nature neuroscience 2021, 24(9):1302–1312.

24. Yang C, Farias F, Ibanez L, Sadler B, Fernandez M-V, Wang F, Bradley J, Eiffert B, Bahena J, Budde J et al: Genomic and multi-tissue proteomic integration for understanding the biology of disease and other complex traits. medRxiv 2020:2020.2006.2025.20140277.

25. Timsina J, Gomez-Fonseca D, Wang L, Do A, Western D, Alvarez I, Aguilar M, Pastor P, Henson RL, Herries E et al: Comparative Analysis of Alzheimer’s Disease Cerebrospinal Fluid Biomarkers Measurement by Multiplex SOMAscan Platform and Immunoassay-Based Approach. Journal of Alzheimer’s disease : JAD 2022, 89(1):193–207.

26. Feng W, Beer JC, Hao Q, Ariyapala IS, Sahajan A, Komarov A, Cha K, Moua M, Qiu X, Xu X et al: NULISA: a proteomic liquid biopsy platform with attomolar sensitivity and high multiplexing. Nature communications 2023, 14(1):7238.

27. Ali M, Archer DB, Gorijala P, Western D, Timsina J, Fernandez MV, Wang TC, Satizabal CL, Yang Q, Beiser AS et al: Large multi-ethnic genetic analyses of amyloid imaging identify new genes for Alzheimer disease. Acta Neuropathol Commun 2023, 11(1):68.

28. Barthelemy NR, Saef B, Li Y, Gordon BA, He Y, Horie K, Stomrud E, Salvado G, Janelidze S, Sato C et al: CSF tau phosphorylation occupancies at T217 and T205 represent improved biomarkers of amyloid and tau pathology in Alzheimer’s disease. Nat Aging 2023, 3(4):391–401.

29. Meeker KL, Butt OH, Gordon BA, Fagan AM, Schindler SE, Morris JC, Benzinger TLS, Ances BM: Cerebrospinal fluid neurofilament light chain is a marker of aging and white matter damage. Neurobiology of disease 2022, 166:105662.

30. Henson RL, Doran E, Christian BT, Handen BL, Klunk WE, Lai F, Lee JH, Rosas HD, Schupf N, Zaman SH et al: Cerebrospinal fluid biomarkers of Alzheimer’s disease in a cohort of adults with Down syndrome. Alzheimer’s & dementia 2020, 12(1):e12057.

31. Zetterberg H, Skillback T, Mattsson N, Trojanowski JQ, Portelius E, Shaw LM, Weiner MW, Blennow K, Alzheimer’s Disease Neuroimaging I: Association of Cerebrospinal Fluid Neurofilament Light Concentration With Alzheimer Disease Progression. JAMA neurology 2016, 73(1):60–67.

32. Eisenstein SA, Boodram RS, Sutphen CL, Lugar HM, Gordon BA, Marshall BA, Urano F, Fagan AM, Hershey T: Plasma Neurofilament Light Chain Levels Are Elevated in Children and Young Adults With Wolfram Syndrome. Frontiers in neuroscience 2022, 16:795317.

33. Crimmins DL, Herries EM, Ohlendorf MF, Brada NA, Garbett NC, Zipfel GJ, Schindler SE, Ladenson JH: Double Monoclonal Immunoassay for Quantifying Human Visinin-Like Protein-1 in CSF. Clin Chem 2017, 63(2):603–604.

34. Sutphen CL, McCue L, Herries EM, Xiong C, Ladenson JH, Holtzman DM, Fagan AM Adni: Longitudinal decreases in multiple cerebrospinal fluid biomarkers of neuronal injury in symptomatic late onset Alzheimer’s disease. Alzheimer’s & dementia : the journal of the Alzheimer’s Association 2018, 14(7):869–879.

35. Willemse EAJ, De Vos A, Herries EM, Andreasson U, Engelborghs S, van der Flier WM, Scheltens P, Crimmins D, Ladenson JH, Vanmechelen E et al: Neurogranin as Cerebrospinal Fluid Biomarker for Alzheimer Disease: An Assay Comparison Study. Clin Chem 2018, 64(6):927–937.

36. Westgard JO, Barry PL, Hunt MR, Groth T: A multi-rule Shewhart chart for quality control in clinical chemistry. Clin Chem 1981, 27(3):493–501.

37. Ostergaard C, Johansen JS, Benfield T, Price PA, Lundgren JD: YKL-40 is elevated in cerebrospinal fluid from patients with purulent meningitis. Clin Diagn Lab Immunol 2002, 9(3):598–604.

38. Deming Y, Filipello F, Cignarella F, Cantoni C, Hsu S, Mikesell R, Li Z, Del-Aguila JL, Dube U, Farias FG et al: The MS4A gene cluster is a key modulator of soluble TREM2 and Alzheimer’s disease risk. Science translational medicine 2019, 11(505).

39. Piccio L, Buonsanti C, Cella M, Tassi I, Schmidt RE, Fenoglio C, Rinker J, 2nd, Naismith RT, Panina-Bordignon P, Passini N et al: Identification of soluble TREM-2 in the cerebrospinal fluid and its association with multiple sclerosis and CNS inflammation. Brain : a journal of neurology 2008, 131(Pt 11):3081–3091.

40. Schindler SE, Cruchaga C, Joseph A, McCue L, Farias FHG, Wilkins CH, Deming Y, Henson RL, Mikesell RJ, Piccio L et al: African Americans Have Differences in CSF Soluble TREM2 and Associated Genetic Variants. Neurol Genet 2021, 7(2):e571.

41. Chatterjee P, Vermunt L, Gordon BA, Pedrini S, Boonkamp L, Armstrong NJ, Xiong C, Singh AK, Li Y, Sohrabi HR et al: Plasma glial fibrillary acidic protein in autosomal dominant Alzheimer’s disease: Associations with Abeta-PET, neurodegeneration, and cognition. Alzheimer’s & dementia : the journal of the Alzheimer’s Association 2023, 19(7):2790–2804.

42. Cruchaga C, Ali M, Shen Y, Do A, Wang L, Western D, Liu M, Beric A, Budde J, Gentsch J et al: Multi-cohort cerebrospinal fluid proteomics identifies robust molecular signatures for asymptomatic and symptomatic Alzheimer’s disease. Res Sq 2024.

43. Cruchaga C, Western D, Timsina J, Wang L, Wang C, Yang C, Ali M, Beric A, Gorijala P, Kohlfeld P et al: Proteogenomic analysis of human cerebrospinal fluid identifies neurologically relevant regulation and informs causal proteins for Alzheimer’s disease. Res Sq 2023.

44. Abe K, Beer JC, Nguyen T, Ariyapala IS, Holmes TH, Feng W, Zhang B, Kuo D, Luo Y, Ma XJ et al: Cross-Platform Comparison of Highly Sensitive Immunoassays for Inflammatory Markers in a COVID-19 Cohort. J Immunol 2024, 212(7):1244–1253.

45. Therriault J, Woo MS, Salvado G, Gobom J, Karikari TK, Janelidze S, Servaes S, Rahmouni N, Tissot C, Ashton NJ et al: Comparison of immunoassay-with mass spectrometry-derived p-tau quantification for the detection of Alzheimer’s disease pathology. Molecular neurodegeneration 2024, 19(1):2.

46. Palmqvist S, Janelidze S, Quiroz YT, Zetterberg H, Lopera F, Stomrud E, Su Y, Chen Y, Serrano GE, Leuzy A et al: Discriminative Accuracy of Plasma Phospho-tau217 for Alzheimer Disease vs Other Neurodegenerative Disorders. JAMA 2020, 324(8):772–781.

