## SupplementaryFigures for "Benchmarking of a multi-biomarker low-volume panel for Alzheimer’s Disease and related dementia research"

**Supplementary Figure 1.** Correlations in CSF for established biomarkers between NULISA and immunoassay measurements for A. Amyloid Beta 40; B. Amyloid Beta 42; C. Amyloid Beta 42/40; D. total tau; E. p-tau181; and F. p-tau217

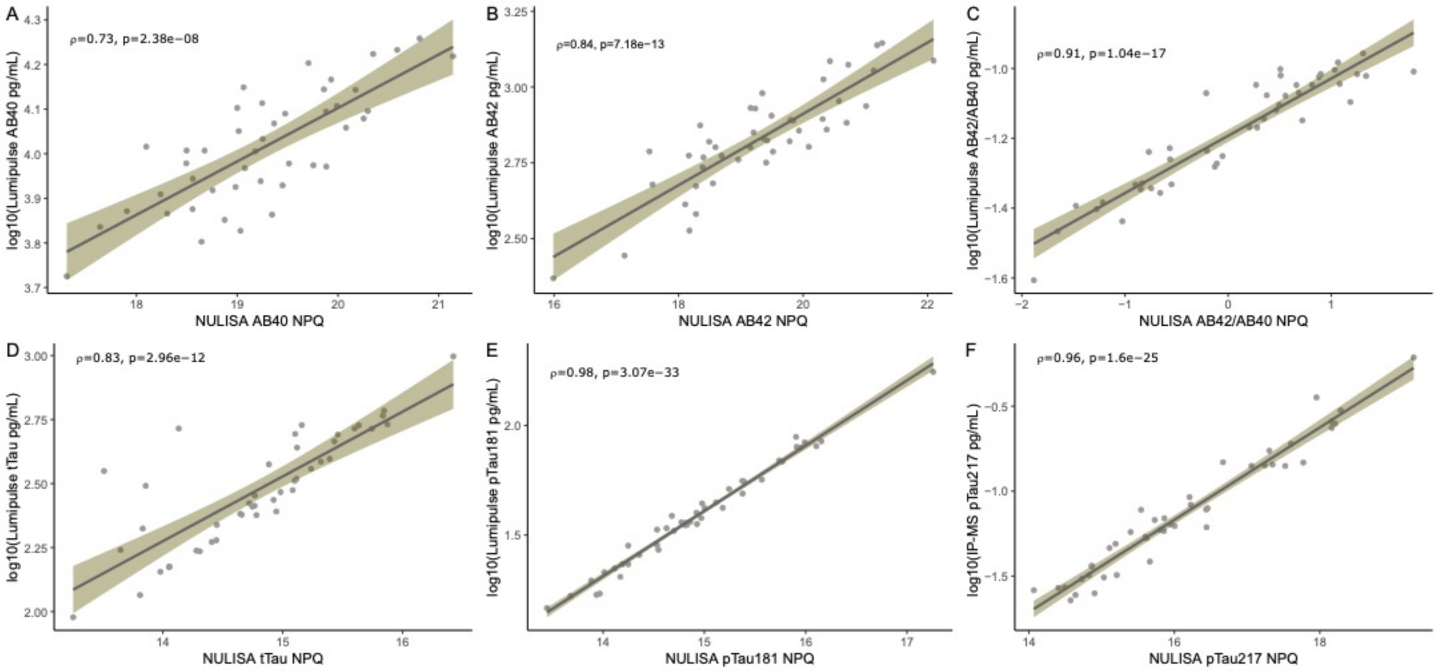

**Supplementary Figure 2.** Correlations values for biomarker measurements and amyloid PET centiloids using A. NULISA Amyloid Beta 42/40 ratio in CSF, B. Lumipulse Amyloid Beta 42/40 in CSF, C. NULISA p-tau217 in CSF, D. IP-MS p-tau217 in CSF; E. NULISA Amyloid Beta 42/40 ratio in plasma, F. IP-MS Amyloid Beta 42/40 ratio in plasma, and G. NULISA p-tau217 in plasma

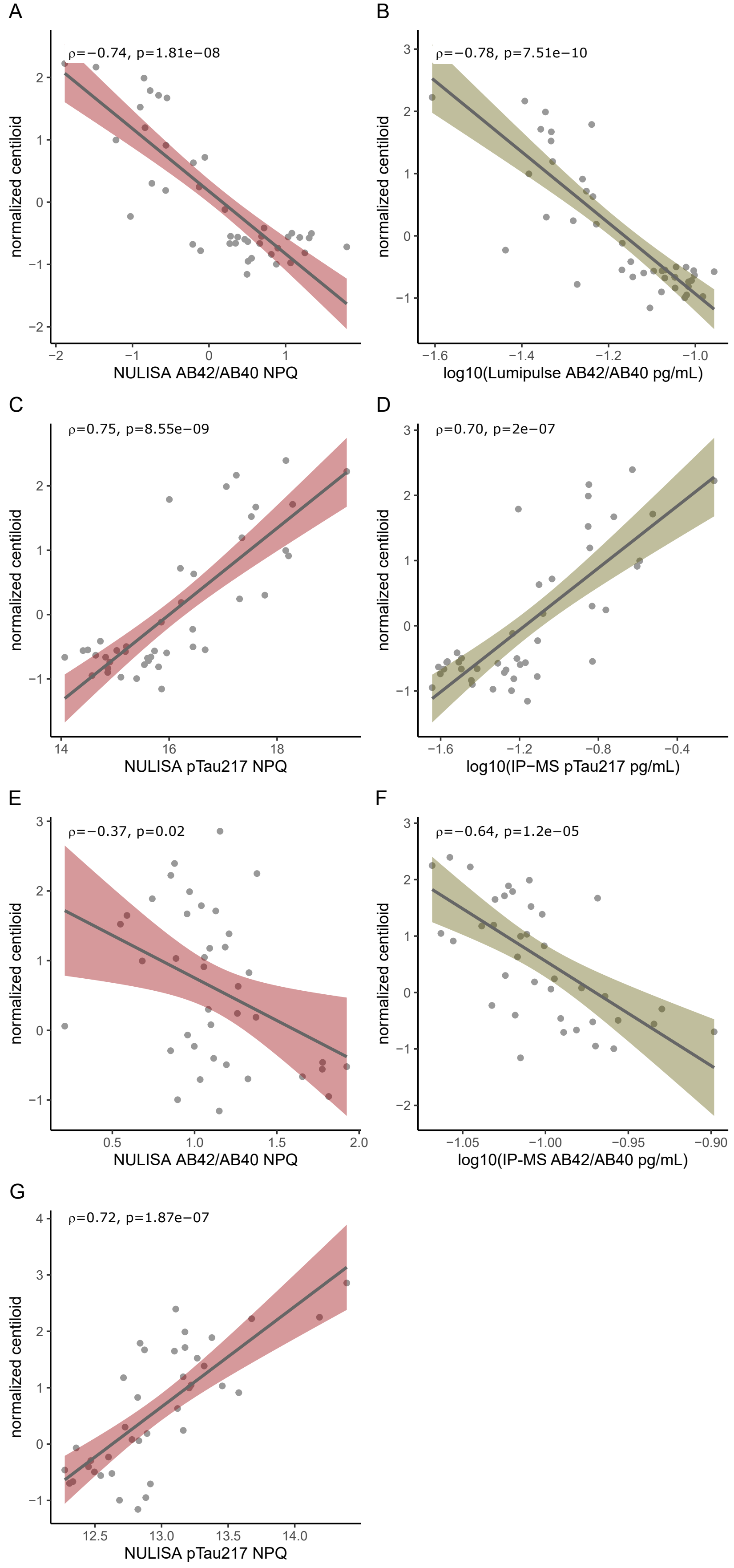

**Supplementary Figure 3.** Correlations in CSF for NFL measurements and NRGN measures assessed with NULUSA and other technologies. Green indicates correlation between NULISA and immunoassay (A, E), blue between NULISA and SomaScan (B, D), and grey correlations between NULISA and Olink (C, F).

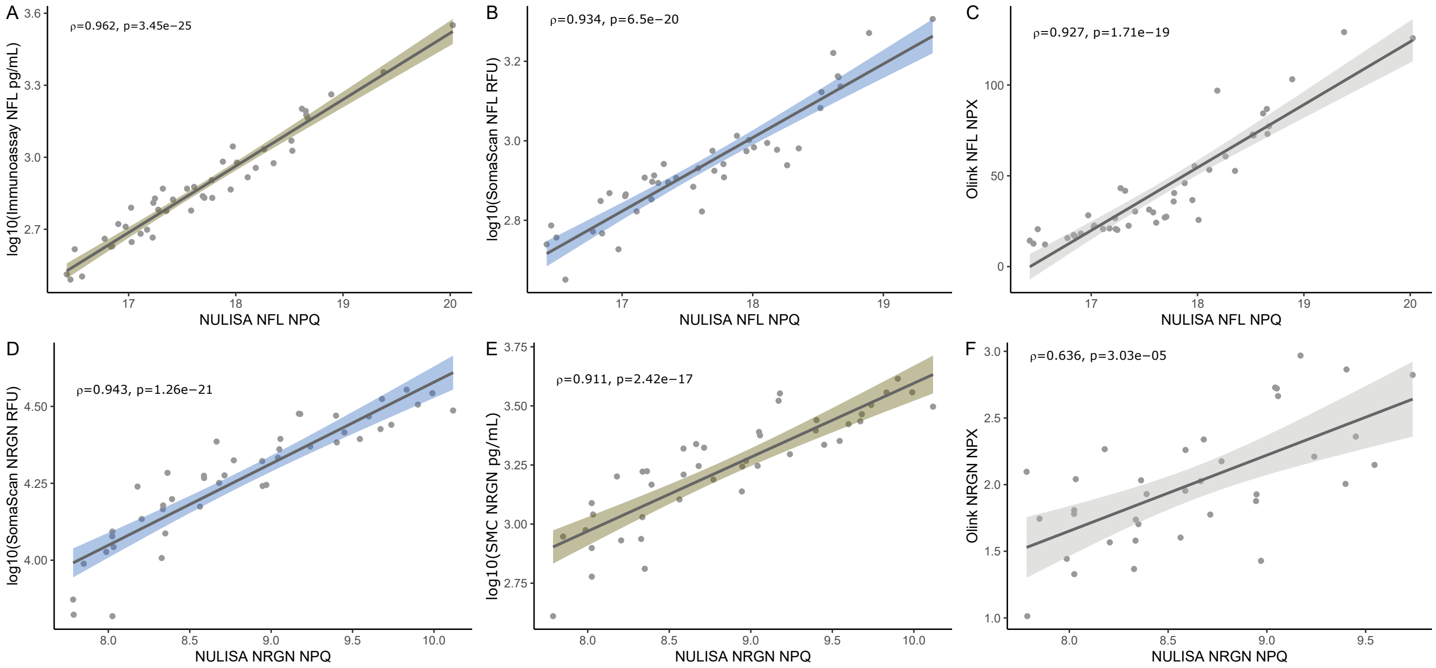

**Supplementary Figure 4.** Correlations in CSF for sTREM2 measurements and YKL40 measures assessed with NULUSA and other technologies. Green indicates correlation between NULISA and immunoassay (A, D), blue between NULISA and SomaScan (B, E), and grey correlations between NULISA and Olink (C, F).

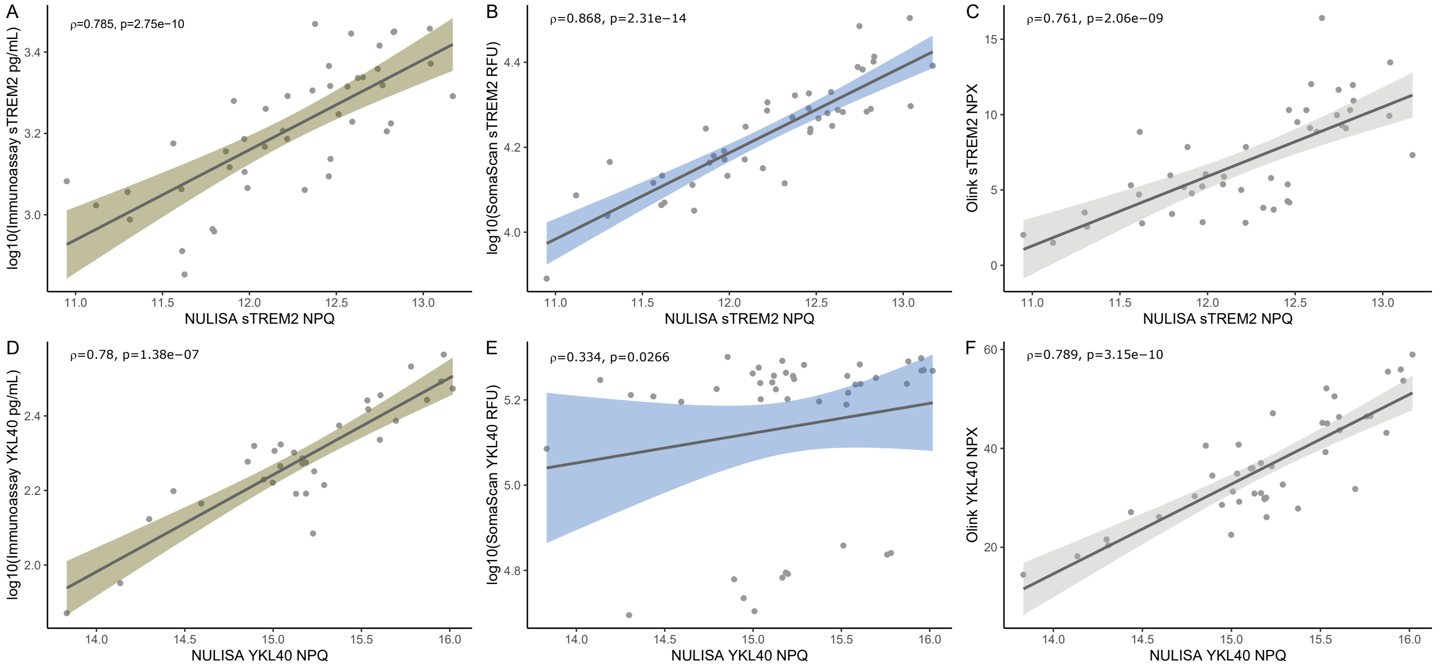

**Supplementary Figure 5.** Correlations in CSF for VILIP1 measurements. SNAP25 and GFAP measures assessed with NULUSA and other technologies. Green indicates correlation between NULISA and immunoassay (A, D), blue between NULISA and SomaScan (B, F, G), and grey correlations between NULISa and Olink (C, E, H).

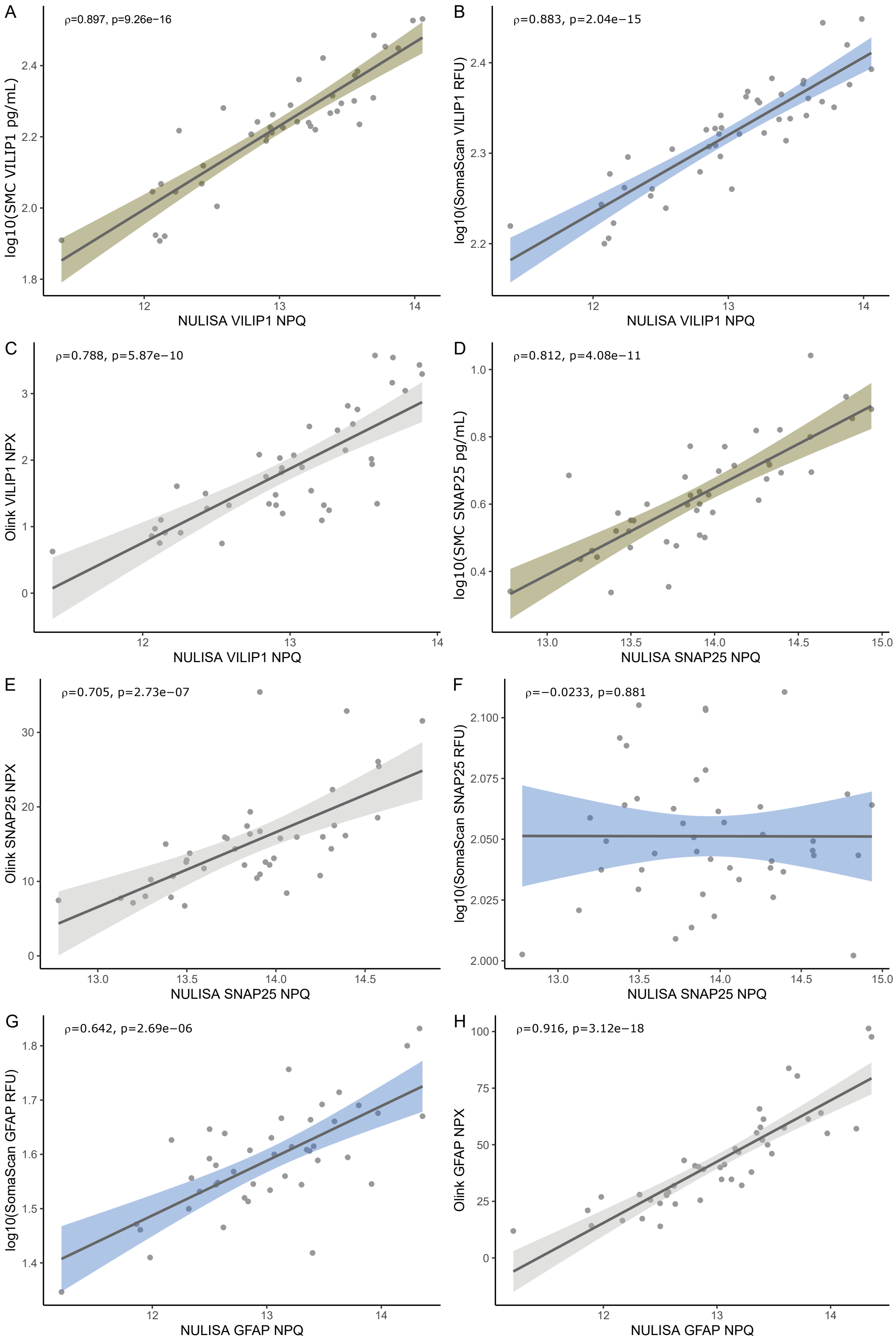

**Supplementary Figure 6** Correlations in Plasma for amyloid beta 40, amyloid beta 42 and its ratio, NFL, sTREM2, NRGN, YKL40, and GFAP. Green indicates correlation between NULISA and IP-MS (A, B, C) or immunoassay (D, I) and blue between NULISA and SomaScan (E, F, G, H).

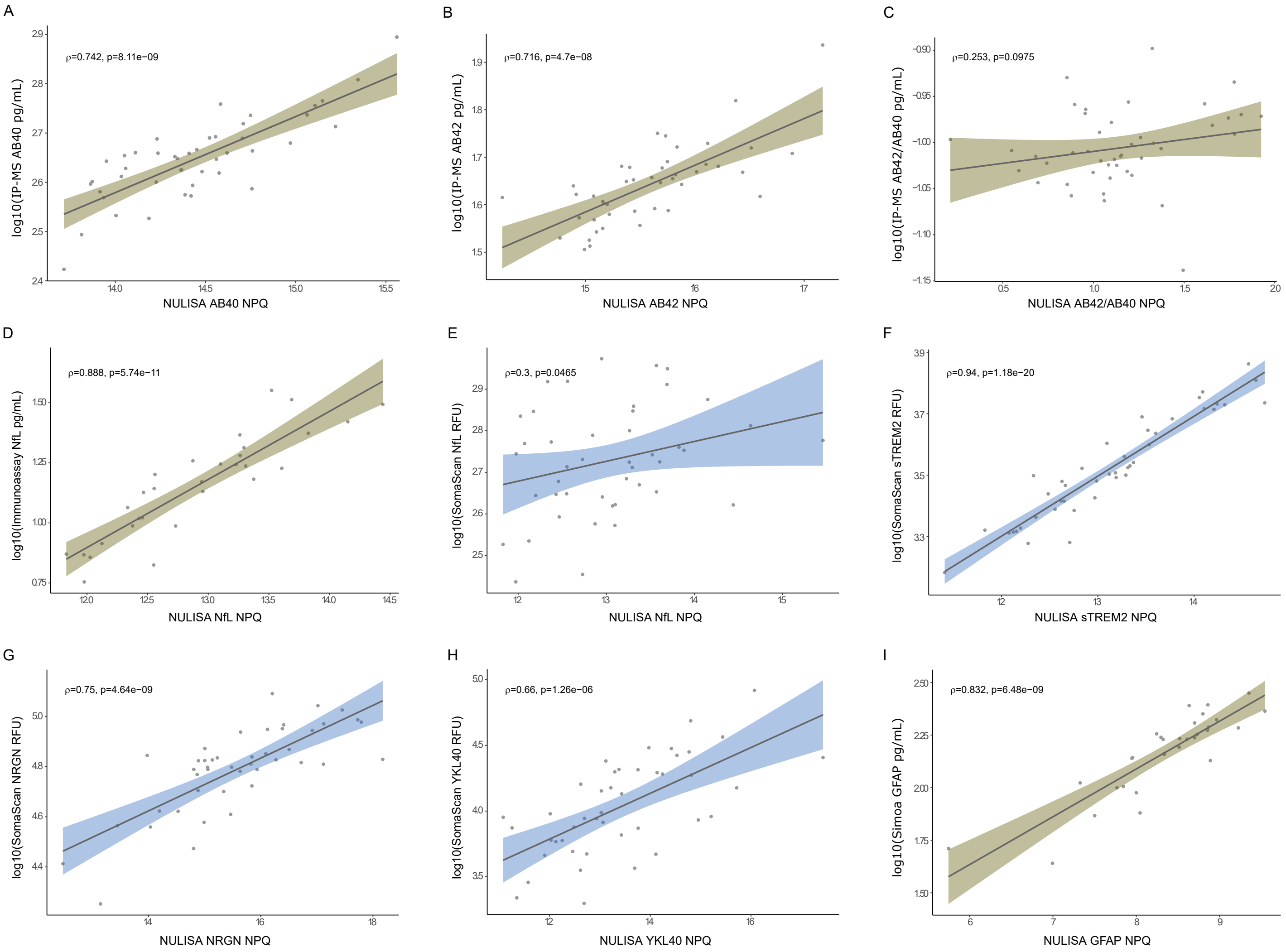

**Supplementary Figure 7.** Correlation values (ρ) distribution in CSF between A. NULISA and SomaLogic, and B. NULISA and Olink and distribution correlation in Plasma between C. NULISA and SomaLogic.

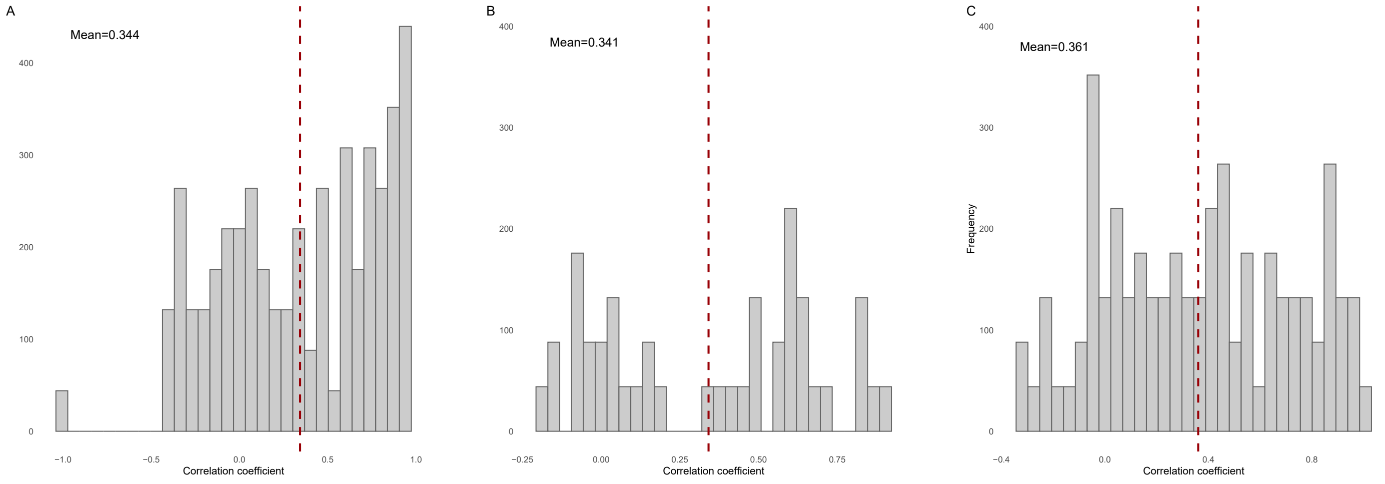

**Supplementary Figure 8.** Volcano plots for the association of the 116 proteins measured using NULISA with Amyloid positivity (based on Pittsburgh Compound-B (PiB) Positron emission tomography (PET) quantifications) in A. CSF; and B. Plasma, and case/control status at draw based on clinical assessment and CDR in C. CSF, and D. Plasma.

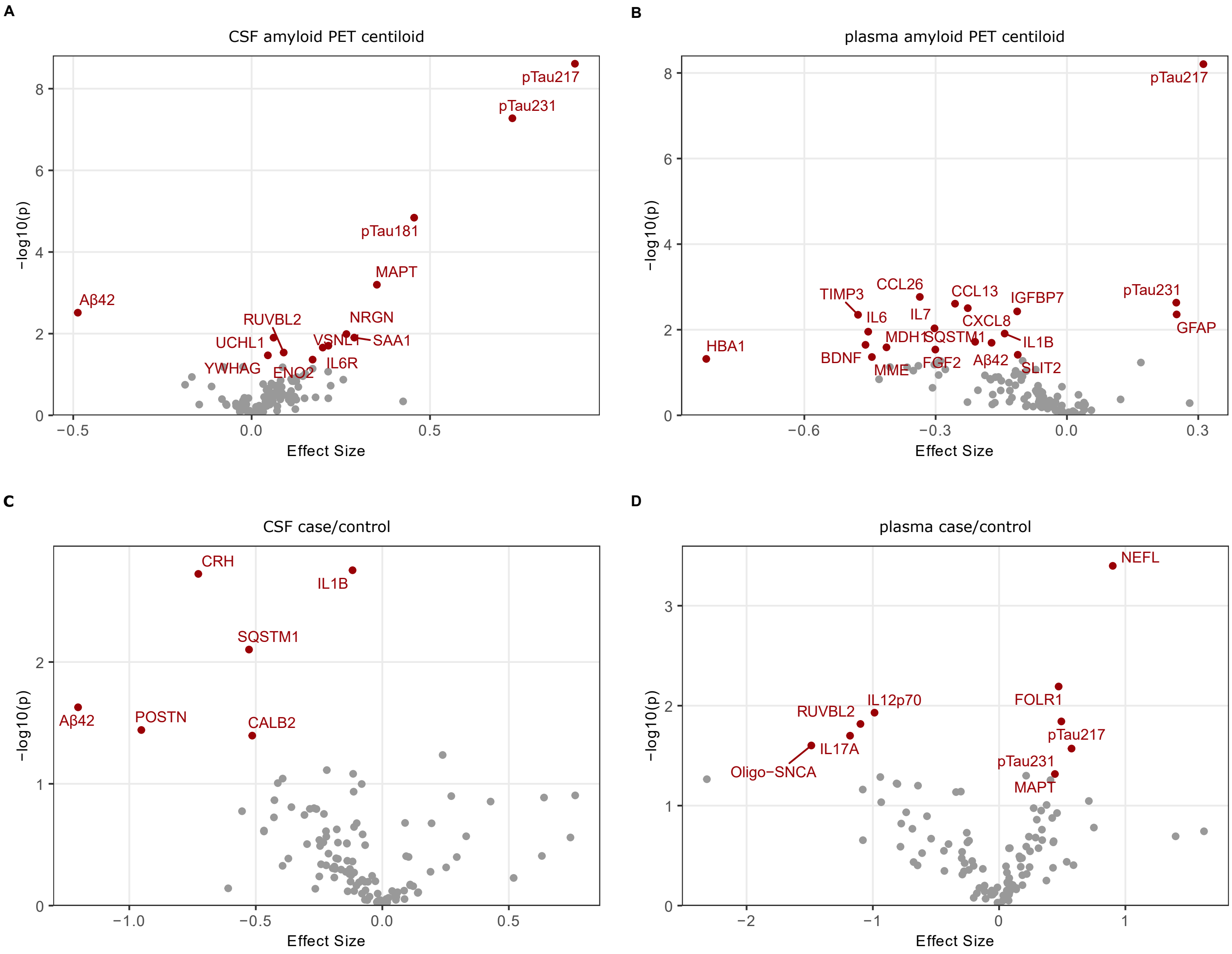
